# Mutations in the tail domain of the neurofilament heavy chain gene increase the risk of amyotrophic lateral sclerosis

**DOI:** 10.1101/2022.11.03.22281905

**Authors:** Heather Marriott, Thomas P. Spargo, Ahmad Al Khleifat, Isabella Fogh, Project MinE ALS Sequencing Consortium, Peter M Andersen, Nazli A. Başak, Johnathan Cooper-Knock, Philippe Corcia, Philippe Couratier, Mamede de Carvalho, Vivian Drory, Jonathan D. Glass, Marc Gotkine, Orla Hardiman, John E. Landers, Russell McLaughlin, Jesús S. Mora Pardina, Karen E. Morrison, Susana Pinto, Monica Povedano, Christopher E. Shaw, Pamela J. Shaw, Vincenzo Silani, Nicola Ticozzi, Philip van Damme, Leonard H. van den Berg, Patrick Vourc’h, Markus Weber, Jan H. Veldink, Richard J. Dobson, Patrick Schwab, Ammar Al-Chalabi, Alfredo Iacoangeli

## Abstract

**Objective:** Genetic variation in the neurofilament heavy chain gene (*NEFH*) has been convincingly linked to the pathogenesis of multiple neurodegenerative diseases, however, the relationship between *NEFH* mutations and ALS susceptibility has not been robustly explored. We therefore wanted to determine if genetic variants in *NEFH* modify ALS risk.

**Methods:** We performed fixed and random effects model meta-analysis of published case-control studies reporting *NEFH* variant frequencies using next-generation sequencing, microarray or PCR-based approaches. Comprehensive screening and rare variant burden analysis of *NEFH* variation in the Project MinE ALS whole-genome sequencing data set was also conducted.

**Results:** We identified 12 case-control studies that reported *NEFH* variant frequencies, for a total of 9,496 samples (4,527 ALS cases and 4,969 controls). Fixed effects meta-analysis found that rare (MAF<1%) missense variants in the tail domain of *NEFH* increase ALS risk (OR 4.56, 95% CI 2.13-9.72, p<0.0001). A total of 591 rare *NEFH* variants, mostly novel (78.2%), were found in the Project MinE dataset (8,903 samples: 6,469 cases and 2,434 controls). Burden analysis showed ultra-rare (MAF <0.1%) pathogenic missense variants in the tail domain are associated with ALS (OR 1.94, 95% CI 0.86-4.37, Madsen-Browning p=0.039), replicating and confirming the meta-analysis finding. High-frequency rare (MAF 0.1-1%) tail in-frame deletions also confer susceptibility to ALS (OR 1.18, 95% CI 0.67-2.07, SKAT-O p=0.03), which supports previous findings.

**Interpretation:** This study shows that *NEFH* tail domain variants are a risk factor of ALS and supports the inclusion of missense and in-frame deletion *NEFH* variants in ALS genetic screening panels.

## INTRODUCTION

Amyotrophic lateral sclerosis (ALS) is a relentlessly progressive and fatal neurodegenerative disease resulting from upper and lower motor neuron loss ^1^. As ALS displays considerable clinical and genetic heterogeneity, it is essential that its genetic mechanisms are defined appropriately in order to develop modifying therapies and enhance personalised medicine approaches ^2^. Around 40-45 genes are implicated in ALS and are involved in cellular processes such as autophagy, DNA damage repair, protein degradation, mitochondrial function and cellular/axonal transport ^3^. The neurofilament heavy chain gene (*NEFH*), encodes the neurofilament heavy subunit protein (NF-H), which regulates several of these activities in an effort to maintain neuronal homeostasis.

Neurofilament protein subunits preserve neuronal architecture by using their side-arms to construct cross-bridges with cytoskeletal components such as microtubules and actin filaments, forming a stable filament-centred matrix that allows intracellular signalling, mitochondrial localisation and ER transport to occur ^4^. This is predominantly orchestrated by the phosphorylation of the head and tail domains of neurofilament genes. For instance, phosphorylation of the head domain acts as a primer for matrix formation, controlling polymerisation of the NF-H subunit in the cell body before the subunits move to the axon, where the lysine-serine-proline (KSP) repeat of the tail domain is phosphorylated to construct the matrix structure and stabilise the neurofilament side arms ^5^. As a result, *NEFH* disruption could influence selective motor neuron degeneration in the brain and spinal cord of affected individuals with ALS via dysregulation of neuronal function ^6^.

Frameshift and missense mutations in *NEFH* have been convincingly linked to the pathogenesis of various neurological diseases, including Charcot-Marie-Tooth disease type 2CC ^7–10^, spinal muscular atrophy ^11^ and Alzheimer’s disease ^12^.

Several lines of evidence suggest hyperphosphorylation of the KSP repeat causes axonal aggregation of phosphorylated NF-H (pNF-H), thereby compromising neuronal integrity and increasing circulating pNF-H levels in the serum and CSF ^5^. Raised pNF-H levels have already been established as a robust biomarker for ALS progression, survival ^13,14^, patterns of motor neuron involvement, and can clinically distinguish ALS from mimics such as hereditary spastic paraplegia, spinal muscular atrophy and myasthenia gravis ^15^. While pNF-H demonstrates prognostic value, there have not been robust studies examining the relationship between *NEFH* mutations and ALS susceptibility. The association between small insertions and deletions in the KSP repeat and ALS risk has been reported in a number of studies ^16–18^, however, this relationship has not been widely reproduced. Still, *NEFH* is included in the majority of genetic screening panels worldwide.

Therefore, a large scale, targeted and comprehensive investigation of the role of common and rare *NEFH* variants in ALS is greatly needed. This study aims to fill such a gap by first performing a meta-analysis of published ALS case-control studies that reported *NEFH* variants and second conducting a large-scale investigation of *NEFH* variation using genetic data from the Project MinE international ALS whole-genome sequencing consortium.

## SUBJECTS/MATERIALS AND METHODS

### Systematic Review

This study was performed in accordance with the 2020 Preferred Reporting Items for Systematic Reviews and Meta-Analyses (PRISMA) guidelines ^19^.

### Eligibility Criteria

Primary research articles published between January 1993 and October 2021 were included if they reported *NEFH* variant frequencies in ALS patients via a candidate or panel gene approach (targeted panel resequencing, variant screening), whole genome sequencing, whole exome sequencing, microarray or PCR-based approaches. Studies were excluded if they were clinical, functional or epidemiological, or if *NEFH* variants were not identified (in targeted gene panel studies) or were identified in non-ALS cases only.

### Information Sources, Search Strategy and Screening Process

Relevant studies were identified by searching PubMed, Embase and Medline databases with the search terms ‘‘amyotrophic lateral sclerosis” OR “ALS’’ in combination with “neurofilament heavy chain gene”, NEFH,” “NFH” OR “NF-H.” After removing duplicate records, title and abstract screening was then performed against the eligibility criteria. Studies which advanced to full text screening were subject to backward citation screening using Web of Science to identify any suitable articles which may have been missed. Full text screening of database and citation identified records was then performed. The search strategy was independently performed, and the results were crosschecked by two members of the team.

### Data Collection Process and Data Synthesis

Once all eligible records were identified, the following study characteristics were extracted; author, publication year, study design, screening method and genetic technology used to detect *NEFH* variants, population (country of origin), study groups, sex and age of ALS groups and diagnostic criteria applied for recruitment into the study. In addition, for each variant identified, the following information was obtained: name according to HGVS protein nomenclature, mutation type, *NEFH* domain location, rsID, and pathogenicity status according to SIFT ^20^ and PolyPhen ^21^ prediction software. Study-specific variant information i.e. frequency in cases and/or controls, odds ratios (ORs) and 95% confidence intervals with p-values and other ALS-associated gene variants carried in *NEFH*-positive individuals, were also extracted. Population-specific *NEFH* variant frequencies were added to each variant record using the gnomAD v2.1.1 non-neuro database ^22^. If the rsID was not supplied, dbSNP ^23^ and gnomAD were searched with the corresponding variant entry. Similarly, for variants without pathogenicity predictions, gnomAD and the hg19 Variant Effect Predictor (VEP) web tool ^24^ was used to obtain variant consequence status.

### Meta-Analysis

Individual missense and exonic insertion and deletion variants which were supported by two or more case-control studies were eligible for variant-level meta-analysis. Subgroup meta-analysis was also performed according to combinations of population-specific gnomAD non-neuro frequency (ultra-rare: < 0.1%, rare: < 1%, or common: > 1%), domain (head, rod, or tail) and variant type. Studies which identified variants which were absent from gnomAD but present in more than one control were classified as common for the stratified analysis. Synonymous variants were excluded from the analysis. Inverse-variance weighted meta-analyses were conducted with both the fixed-effect (Cochran-Mantel-Haenszel) and random-effect (DerSimonian-Laird) models. Crude ORs were calculated from the extracted data. Between-study heterogeneity was assessed using the combination of the I^2^ test and Cochran-Q statistic, with significant heterogeneity indicated when I^2^ > 50% and Q < 0.10. Publication bias was assessed with both Egger’s and Harbord’s test, with p-values < 0.05 classed as displaying significant outcome heterogeneity and selective reporting. All statistical analyses were performed using the *metabin* and *metabias* functions of the meta R package ^25^.

### Genetic Screening

Whole-genome sequencing samples collected as part of the Project MinE ALS sequencing consortium ^26^ were used to investigate the impact of *NEFH* variants in ALS and for replicating the literature based meta-analysis results. Sample overlap between Project MinE and the studies included in the meta-analysis was investigated by contacting the authors of the studies that included patients of the same nationality as the Project MinE participants. No overlap was found. No Briefly, genomic DNA from venous blood was isolated using standard methods and assessed with gel electrophoresis before PCR-free 100bp paired-end sequencing was performed on the Illumina HiSeq2000 platform, which yielded ∼40x coverage. The full dataset consists of 9,050 samples, which includes 6,603 individuals defined as having pure ALS as well as 2,447 age and sex matched controls. After standard quality control measures, the final filtered dataset comprised of 6,469 ALS cases and 2,434 controls from 13 countries (Supplementary Table 1) for which whole-genome SNV and small indel data were available ^27^. Structural variant data generated with Manta ^28^ were available for approximately two thirds of samples (4,686 ALS cases and 1,859 controls). Variants were aligned to hg19. Variants were then annotated with VEP for both functional consequence/type (i.e. UTR, intronic, missense, indel, synonymous) and VEP-specific classification of the impact of the variant (i.e. high, moderate, low, modifier). Screening of structural variants called with Manta v0.28.0 ^28^ was also performed; SURVIVOR v1.0.9 ^29^ was used to create a union callset before variants greater than 100,000bp were excluded to reduce false positives. The remaining variants were then annotated with AnnotSV ^30^ and CADD-SV ^31^ to assess their potential pathogenicity. All results files were converted into a matrix with the VariantAnnotation R package ^32^, from which case-control frequencies were calculated. For the review-identified variants and structural variants present in Project MinE, Firth logistic regression was performed using RVTests ^33^ under default settings to assess potential associations between variant status and ALS susceptibility. Results were corrected for sex and the first 10 principal components.

### Rare Variant Burden Analysis

Burden analysis of all *NEFH* variants identified in the Project MinE samples was performed with RVTests ^33^, using Madsen-Browning and SKAT-O methods under default settings. Results were corrected for sex and the first 10 principal components. Variants were initially grouped by frequency (ultra-rare: <0.1%, high-frequency rare: 0.1-1%), according to the highest value in control databases (gnomAD non-neuro non-Finnish European and Project MinE controls), before being grouped by functional domain (whole gene, head, rod, tail) for which the genomic coordinates were obtained with the *ensembldb* R package. For each functional domain, variant burden was calculated for several variant types (missense, synonymous, insertion, deletion, 3’ UTR, 5’ UTR, intronic) and VEP impact classes (high, moderate, low, modifier). Variant burden was also repeated, sub-setting missense variants into predicted pathogenicity classes, according to SIFT and/or Polyphen scores (“deleterious,” “deleterious low confidence,” “possibly damaging” or “probably damaging”).

## RESULTS

### Study Selection

The literature systematic review process flowchart is presented in Figure 1. The initial search identified 29 articles which were eligible for title and abstract screening, of which 16 articles were the wrong study type, disease, or instances where genetic screening did not include *NEFH* or identify *NEFH* variants even if *NEFH* was present in the targeted sequencing panel. Backward citation searching of the remaining 13 articles found an additional 251 records for screening. Manual full text inspection removed a further 242 records (2 from database search and 240 from citation search) as the inclusion criteria were not met. In total, 22 studies involving a total of 10,959 individuals (6,090 ALS cases and 4,869 controls) from 14 countries were included in the present study.

**Figure 1.**
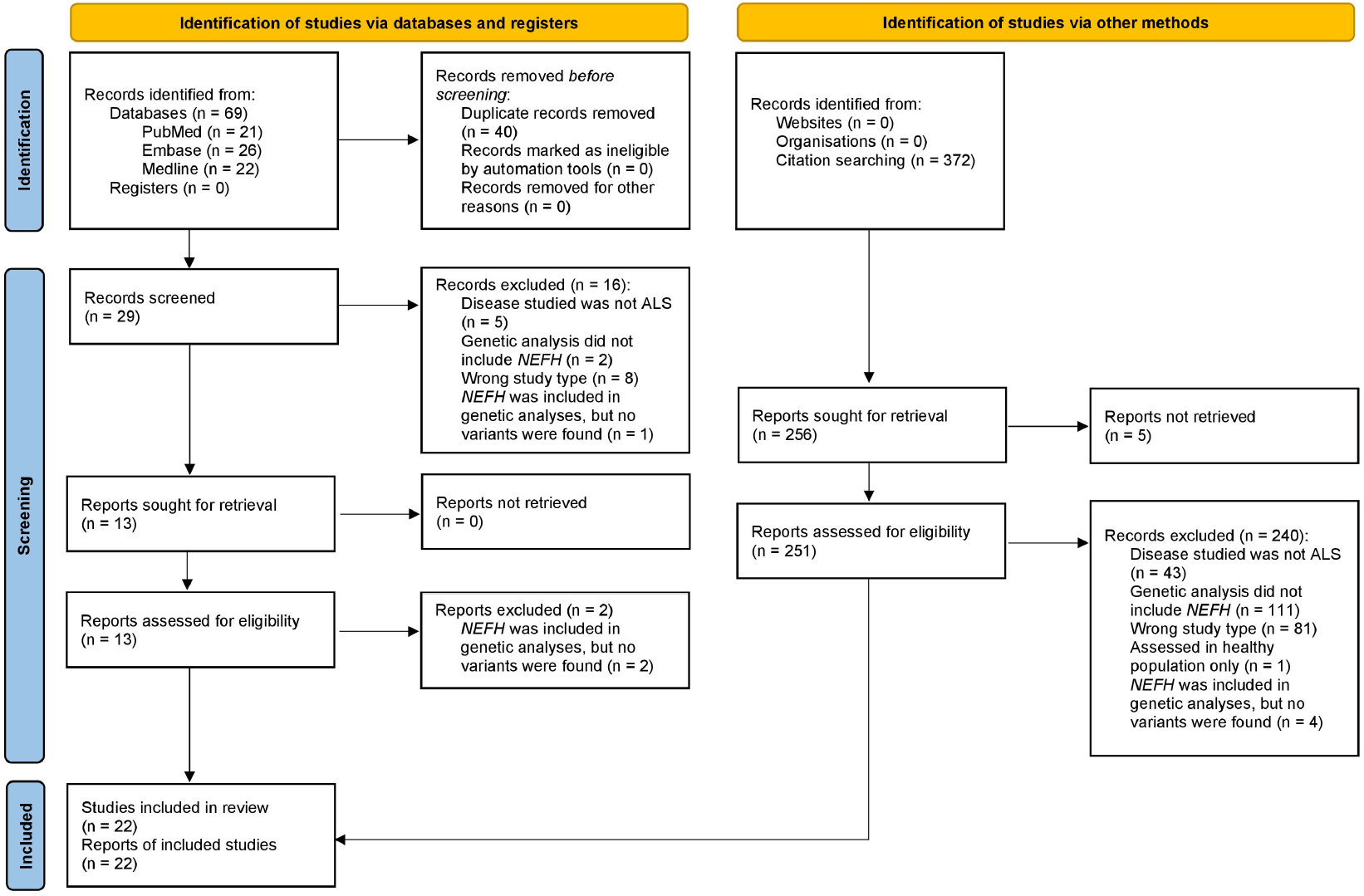
PRISMA flowchart of the study systematic review process. The left of the figure outlines screening for articles identified via PubMed, Embase and Medline databases, whilst the right outlines the process for articles found via backwards citation screening of articles undergoing full-text screening.

### Study Characteristics

An overview of the characteristics of all included studies is given in Table 1. The people were most frequently sampled from Asian (N Studies = 7) and European (N Studies = 7) populations, with a family disease history reported in 77% of studies. Diagnostic criteria were applied to support inclusion in 15 studies (68%), with varying definitions of El Escorial criteria employed in 93% of those. A combination of El Escorial and Awaji-Shima criteria was used in one study. The average age of recruitment of the ALS patients ranged from 30.7 to 62.1 (median 58.1), with a male: female ratio ranging between 0.60 and 1.78 (median 1.38) across studies. When separating by country, Asian populations had a younger median age at recruitment and a higher median male: female ratio than European populations (Asian: age 52.01, sex ratio 1.52; European: age 60.1, sex ratio 1.22). A case-control design was adopted in 12 studies (55%), with 6 investigating *NEFH* variation in ALS via candidate gene-based methods. Gene panels which included *NEFH* were used in 13 studies, with a further 2 opting for custom variant panel screening. The most popular genetic technology used to identify *NEFH* variants was whole-exome sequencing (N Studies = 6) and the combination of whole-exome sequencing with validation approaches such as PCR and Sanger sequencing (N Studies = 6).

**Table 1.**
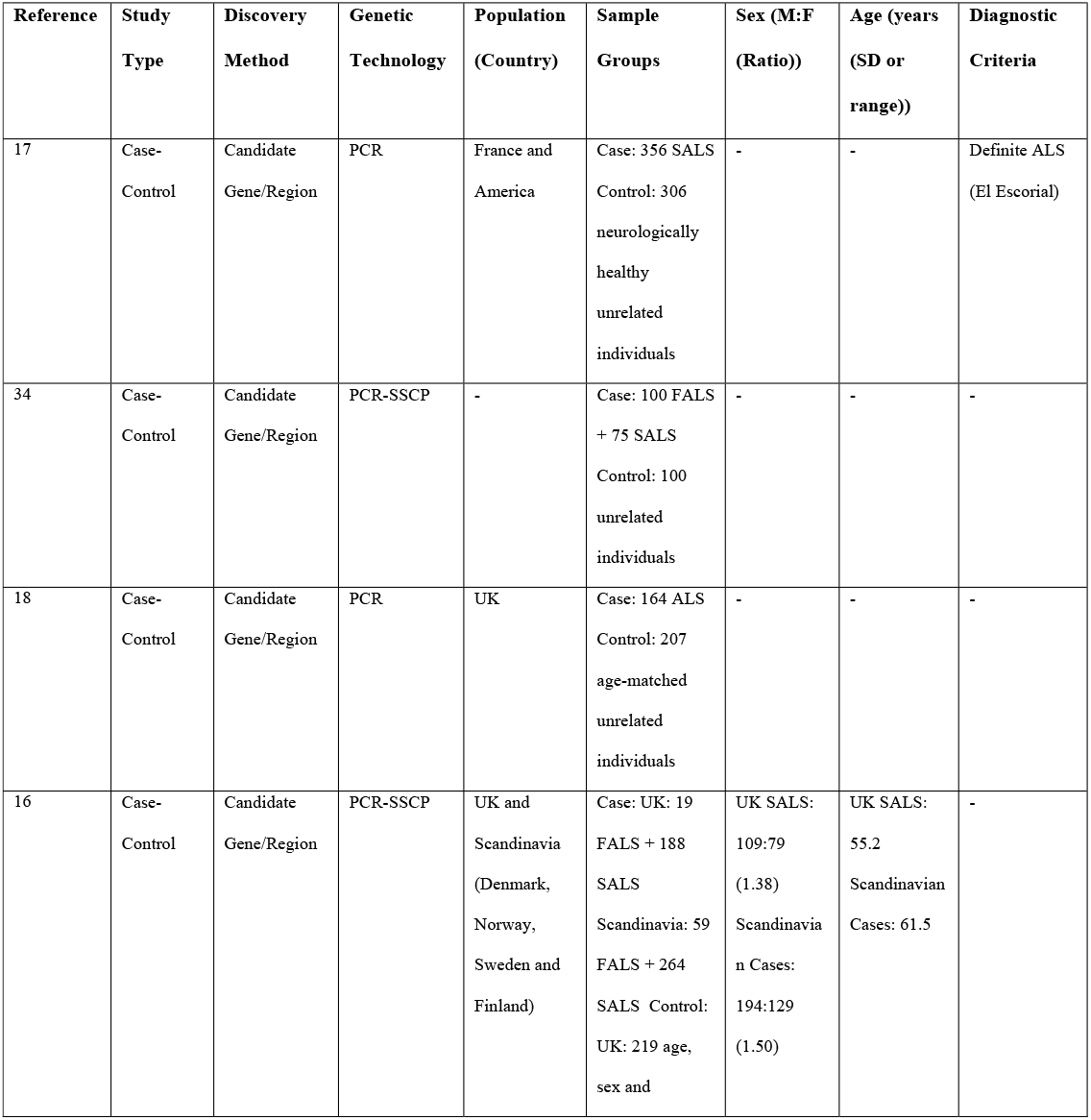
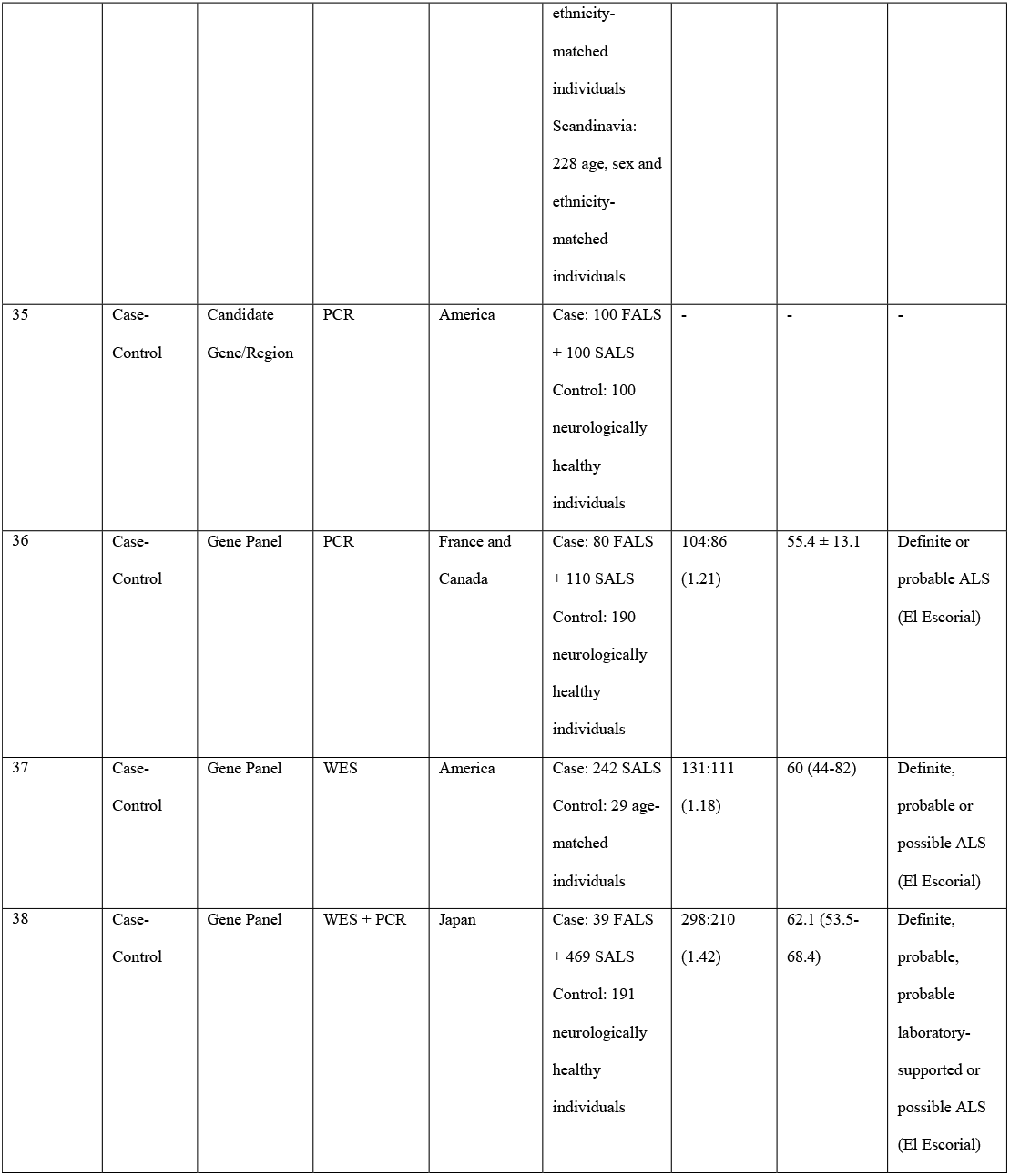
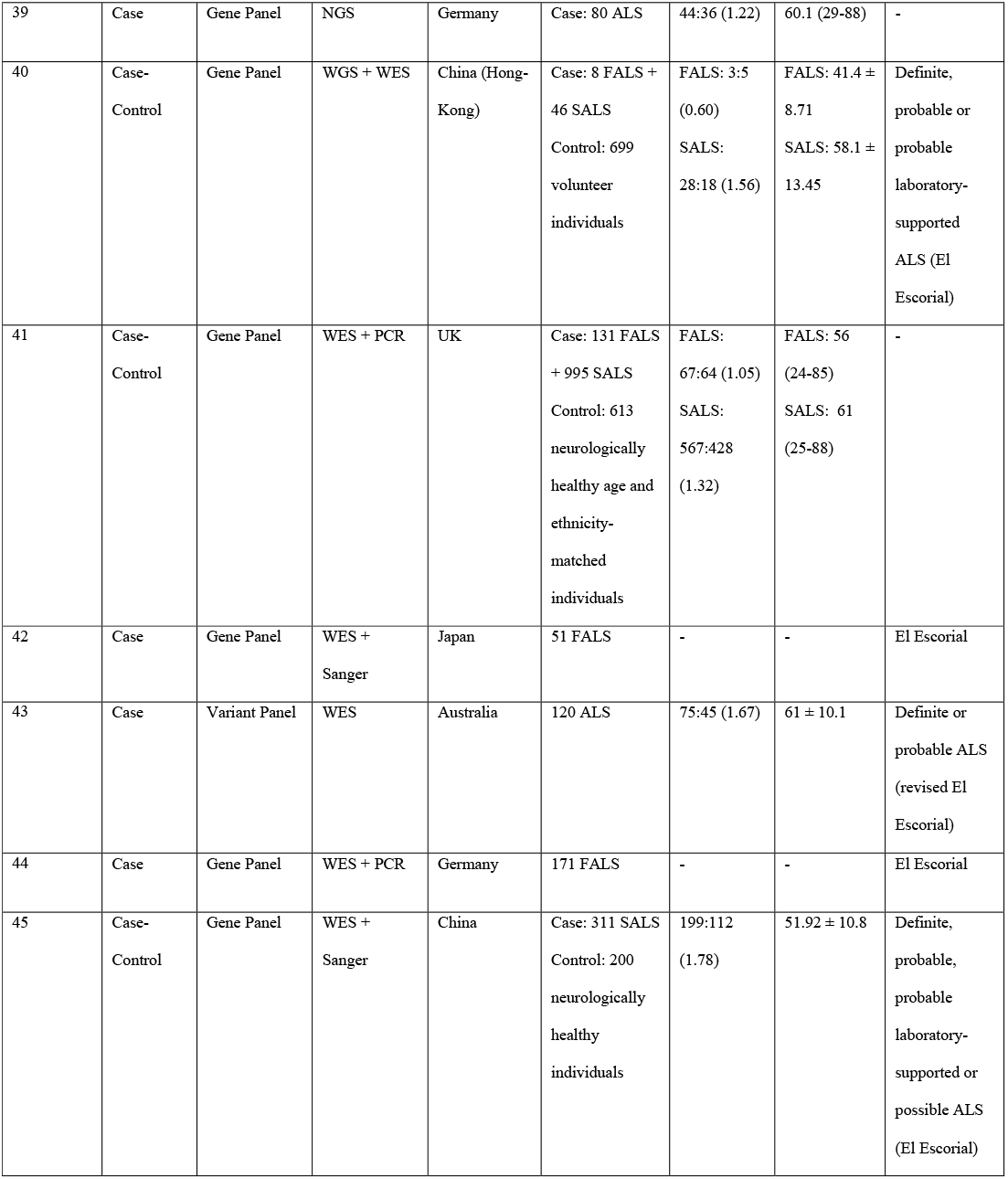
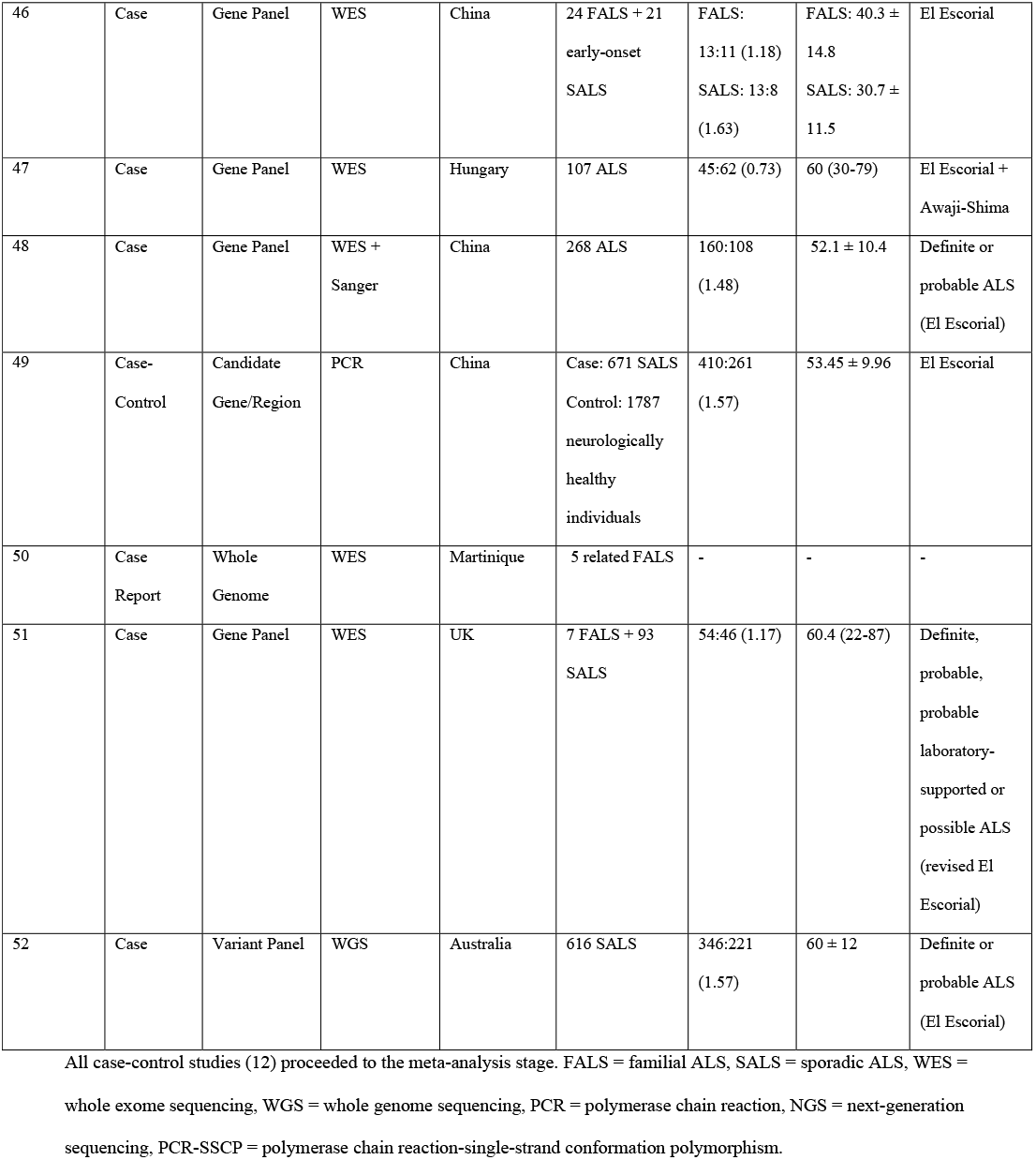
Summary characteristics of all included studies identified from the systematic review.

### Variant Characteristics

A total of 59 *NEFH* variants were identified from the included studies. Full details of each variant are documented in Supplementary Table 2, with their genomic coordinates and base pair substitutions (for hg19) available in Supplementary Table 3. Missense variants were the most represented (67.8%), followed by inframe deletions (13.6%), synonymous variants (13.6%), inframe insertions (1.7%), frameshift deletions (1.7%) and stop-gained SNVs (1.7%). Insertion/deletion variants (indels) ranged from 3bp to 48bp in length and exclusively occupied the tail (Figure 2). Seven times as many in-frame deletions were found in the KSP repeat than the lysine-glutamic acid-proline (KEP) segment. Only 2 variants were found in the head domain (Figure 2). Only 17 variants (28.8%) were reported in more than one study. Ten people with *NEFH* variants also harboured variants in other ALS-associated genes, including *SOD1, FUS, OPTN, SETX, ALS2* and *CHMP2B* (Supplementary Table 2).

**Figure 2.**
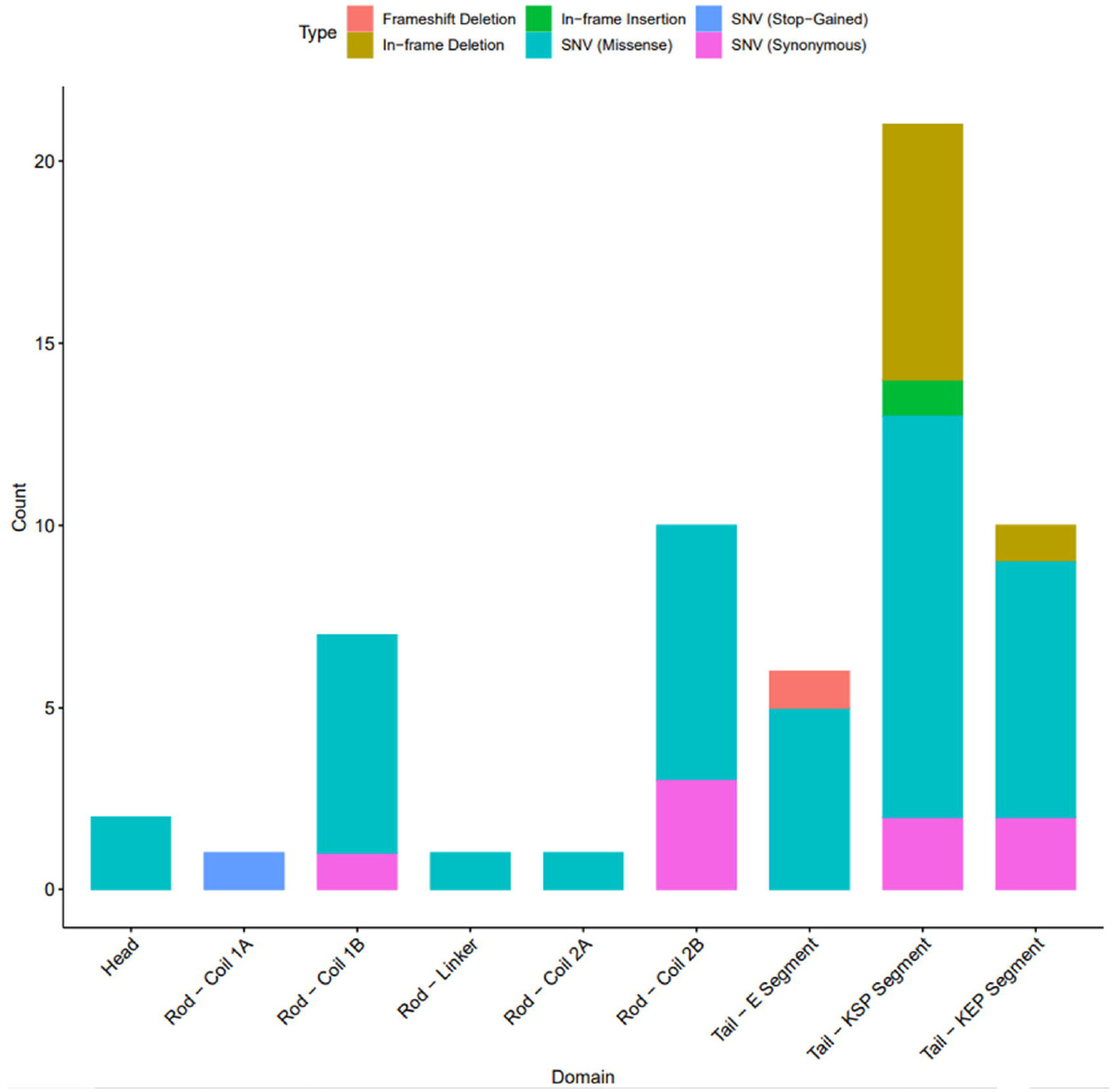
*NEFH* domain distribution of the 59 variants identified from the systematic review. Colours characterise the different variant types. KEP = lysine-glutamic acid-proline; KSP = lysine-serine-proline.

### Meta-Analysis of Previously Published Studies

The twelve case-control studies we identified were selected for meta-analysis. A total of 34 deletion, insertion and missense variants were reported across these studies (displayed in the top panel of Figure 3) in a total of 9,496 individuals (4,527 cases; 4,969 controls). Of these, 8 variants (3 in-frame deletions and 5 missense) were identified in two or more case-control studies and therefore were included in the variant-level meta-analysis. No singular variant was shown to significantly confer or reduce risk for ALS (Supplementary Table 4). One of the deletion variants, K790del, displayed a significantly high level of heterogeneity according to both Cochran’s Q and I^2^ metrics (Supplementary Table 4).

**Figure 3.**
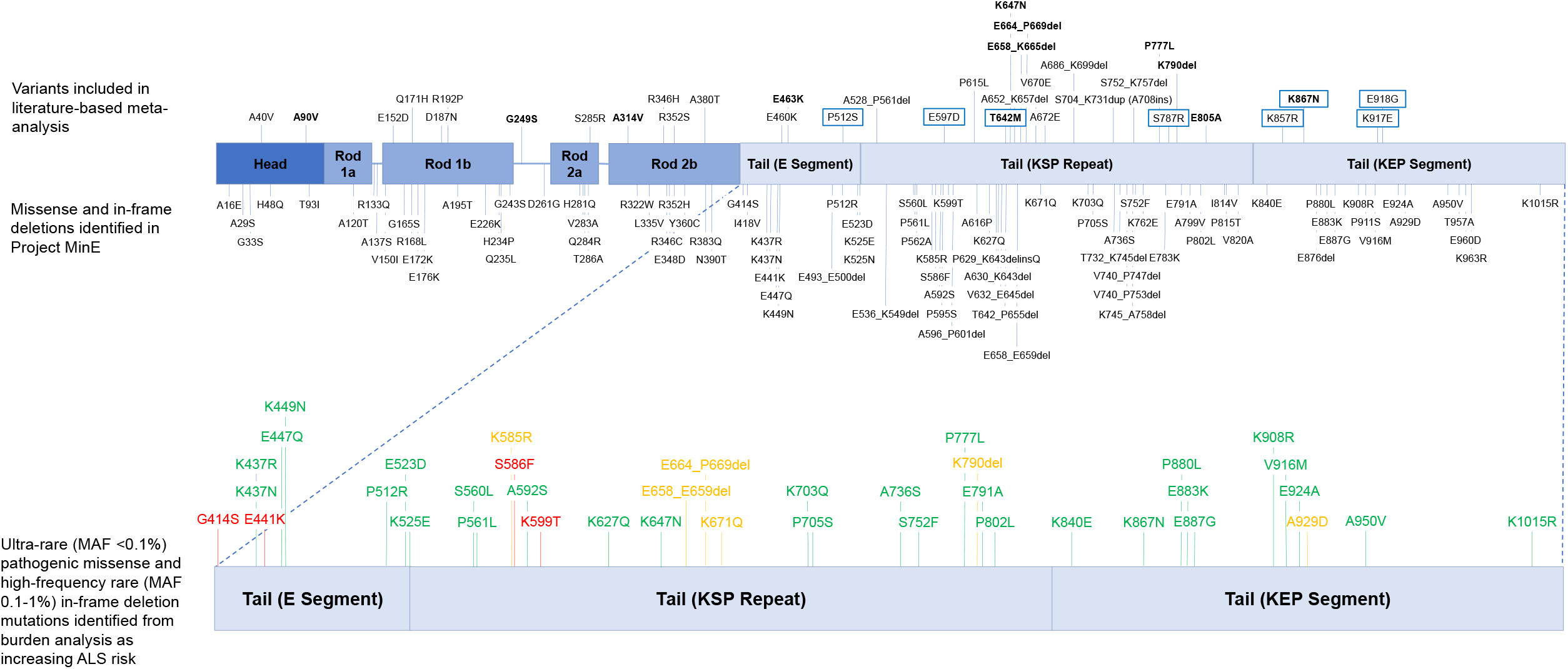
Schematic depicting the locations of the gene variants included in the meta-analysis and in both meta-analysis and Project MinE (top), as well as the variants that were found to increase the risk for ALS with burden analysis (bottom). Green = only present in cases. Amber = present in cases and controls. Red = only present in controls.

We performed meta-analyses of *NEFH* variants based on the aggregation of variants stratified by frequency, domain and variant type (see methods). We found that rare missense variants in the tail domain increase the risk of ALS (Table 2, Figure 4), with an OR of 4.56 (95% CI 2.13-9.72, p<0.0001) under the fixed-effects model. There was no evidence of inter-study heterogeneity (Cochran’s Q = 2.30, p=0.51, I^2^ = 0%) or publication bias (Egger p=2.11, Harbord p=1.85). We also found that rare missense and rare tail variants were also associated with an increased risk of ALS (Table 2), with ORs of 2.37 (95% CI = 1.39-4.04, p=0.0015) and 2.42 (95% CI = 1.28-4.58, p=0.0066), although we determined that the rare missense tail variants were driving this result, as removing these from the meta-analyses caused both associations to be lost. Across all categories, deletion variants did not significantly increase or reduce susceptibility for ALS (Table 2).

**Table 2.**
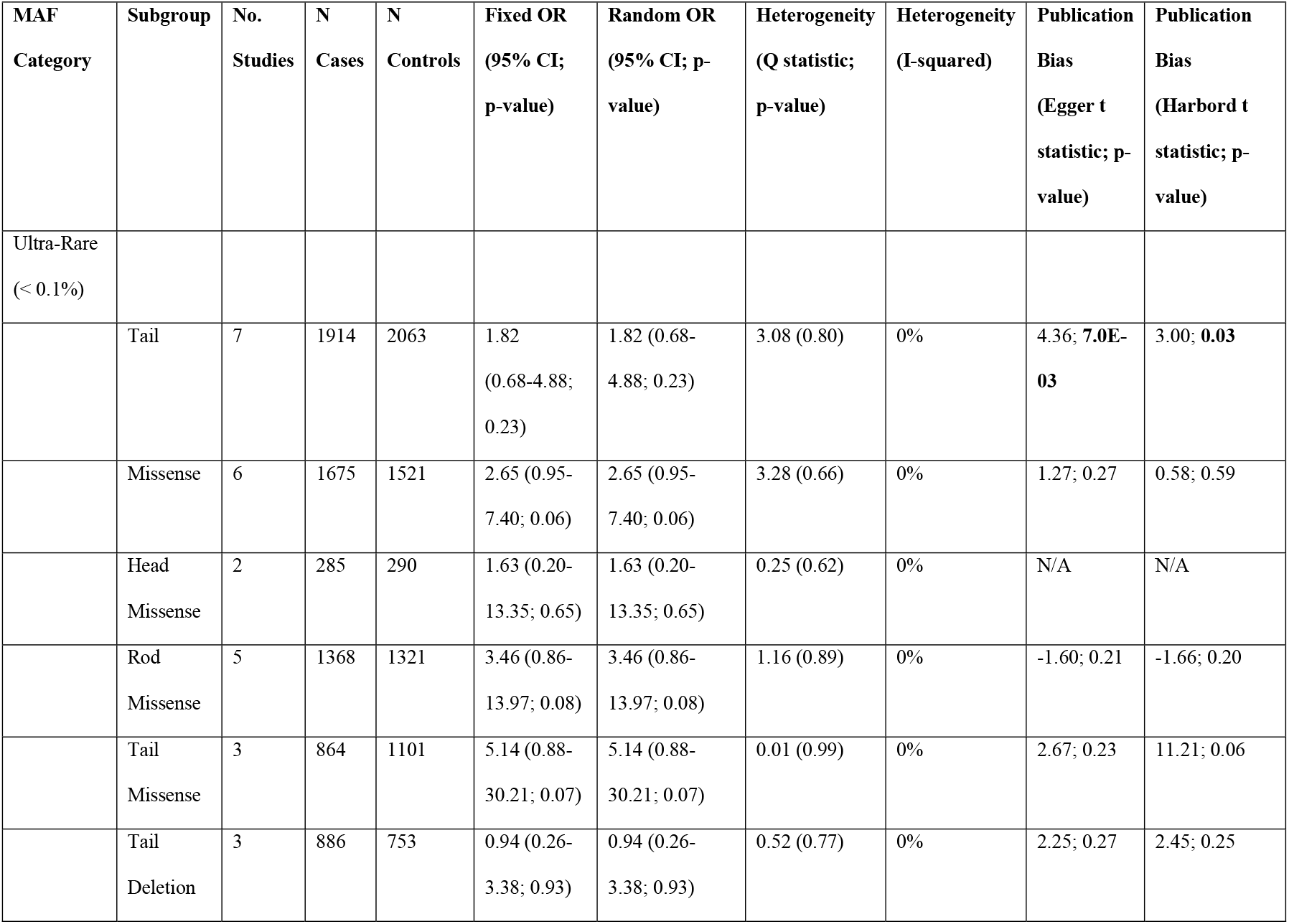
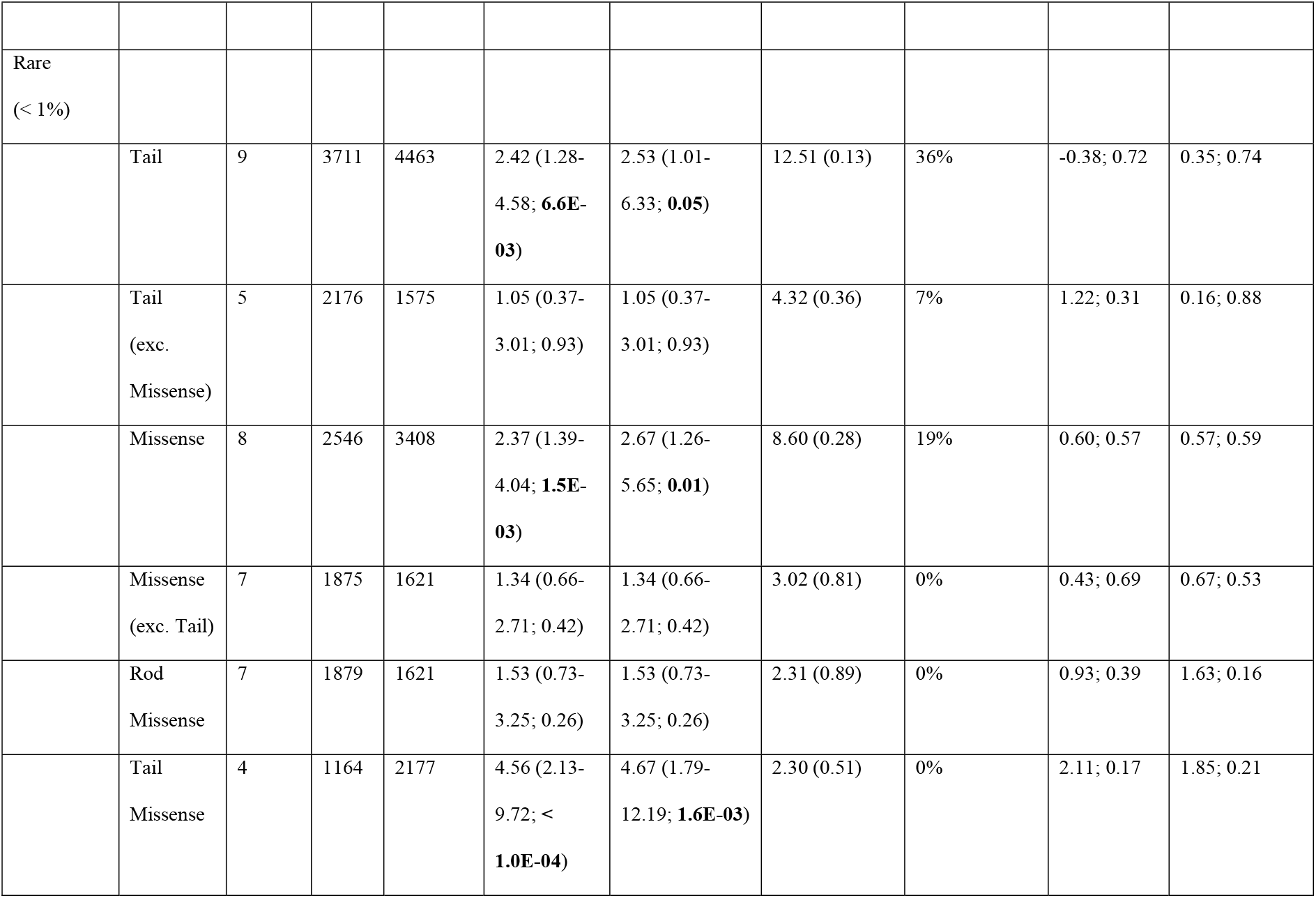
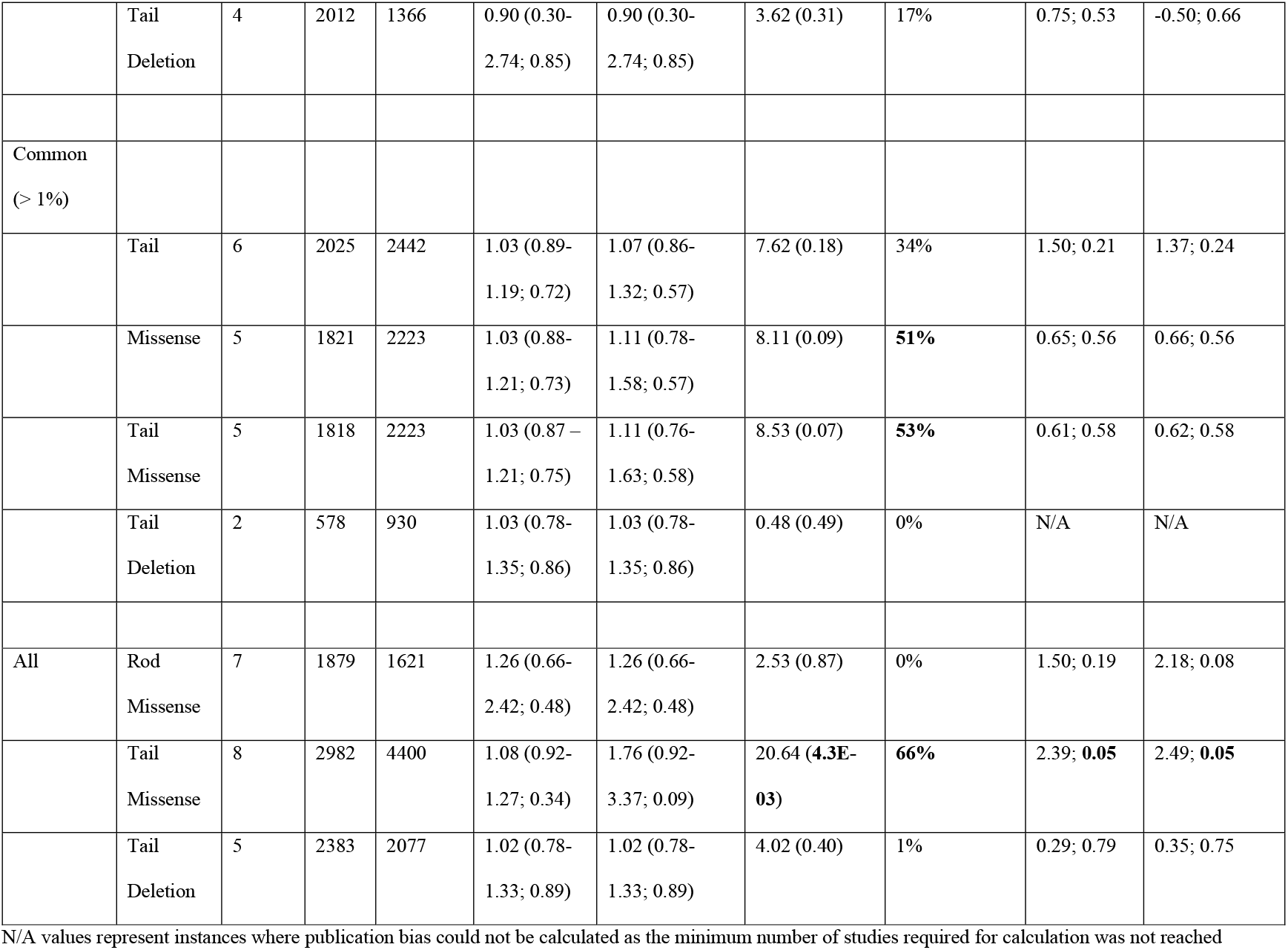
Results of the subgroup meta-analysis.

**Figure 4.**
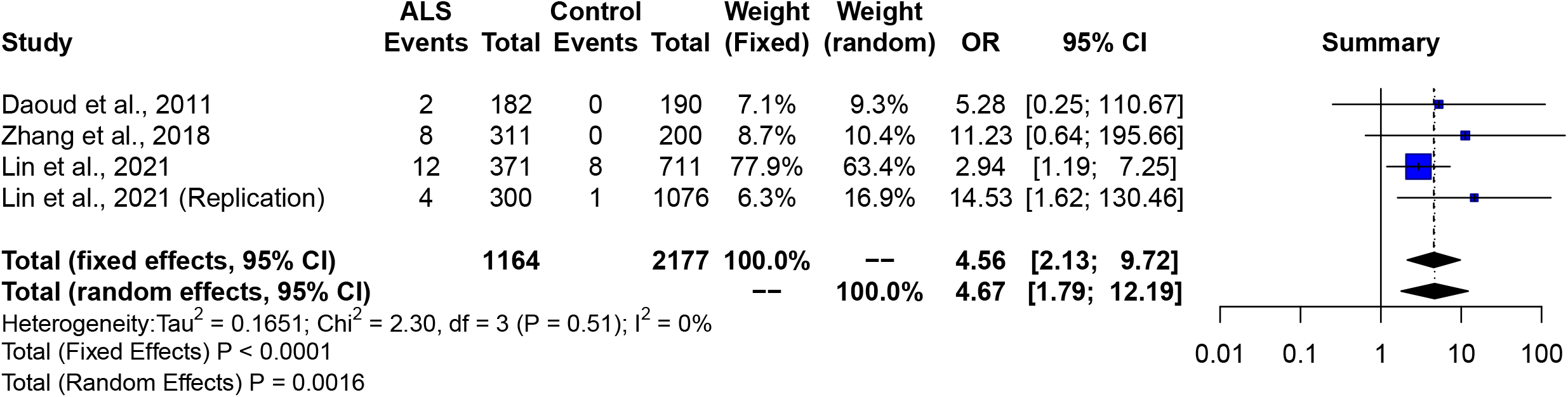
Forest plot demonstrating that rare (MAF<1%) missense variants in the tail domain increase the risk of ALS.

### Screening of SNV/indel *NEFH* variants in the Project MinE cohort

We next screened the whole *NEFH* gene in the Project MinE dataset (6,469 ALS cases and 2,434 controls) to obtain a complete landscape of *NEFH* variation in ALS. A total of 591 SNV and indel variants were identified (Figure 5a). Additional information on all variants are available in Supplementary Table 5. The KSP repeat harboured the highest number of variants (61; 10.32%). Interestingly, intronic regions contained 65% of all variants found in the cohort, with 220 (57.29%) existing as singletons (in either one case or one control). In fact, there were 351 singletons totalling 59.39% of all variants identified in the cohort (Figure 5b), with 220 (62.68%) being in intronic regions of *NEFH*; 3.4x that of the tail domain (65 variants; 18.52%). When accounting for variants from different sources, 462 (78.17%) were identified only in cases and/or controls and not the review or gnomAD non-neuro non-Finnish database (Figure 5c), and are therefore classified as ‘novel’ for the purposes of this study.

**Figure 5.**
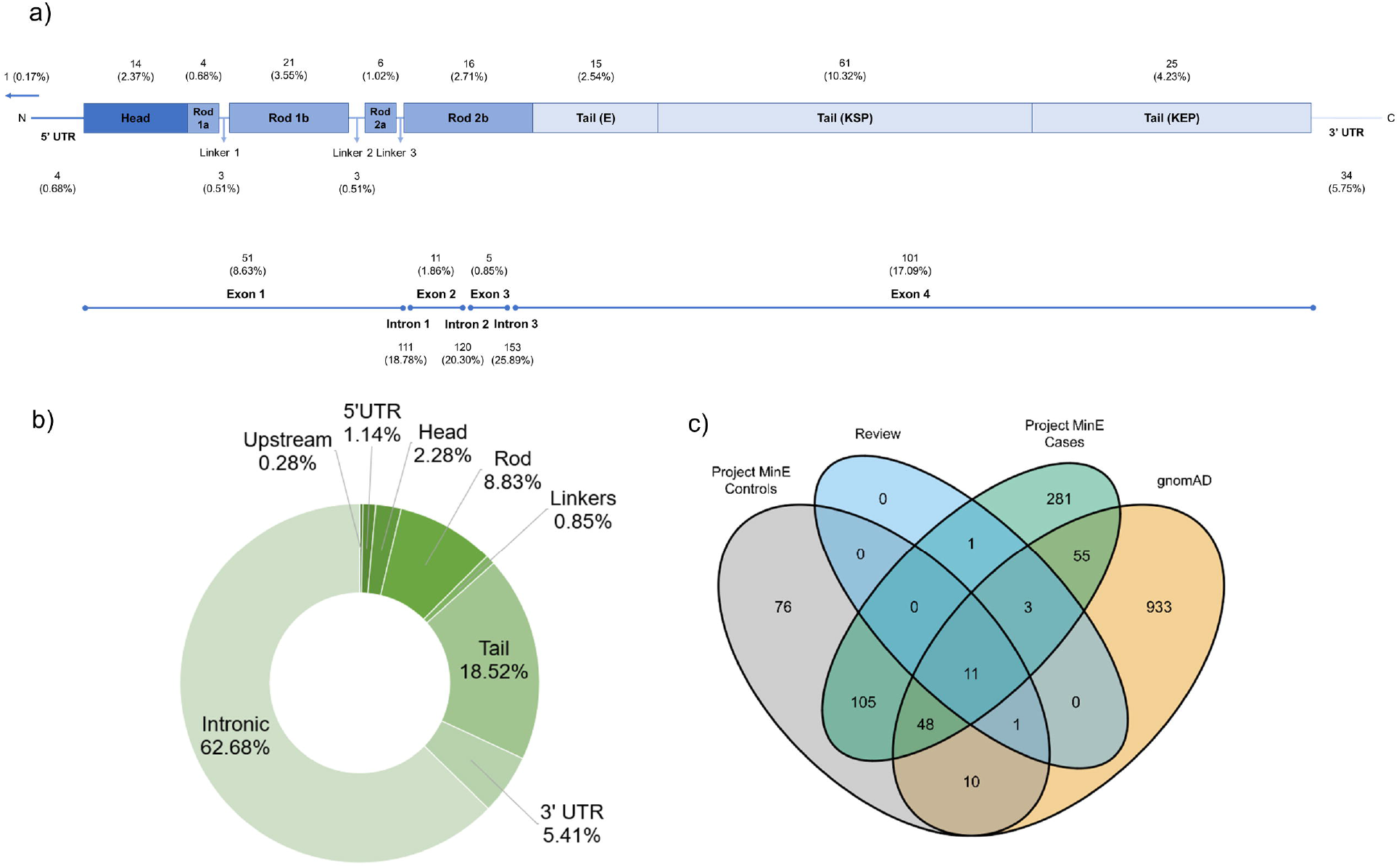
Results of the SNV/indel screening analysis in the Project MinE cohort. Additional information on all 591 variants identified are available in the Supplementary Information. a) Proportion of variants found in various gene domains and untranslated regions (top), and in exons and introns (bottom). b) Breakdown of the 351 *NEFH* singletons by domain. c) A Venn diagram illustrating the overlap of the *NEFH* variants in Project MinE cases and controls, the systematic review and the gnomAD v2.1.1 database. The value for variants that are only in gnomAD (933) refers to the remaining *NEFH* variants in the catalogue after accounting for variants shared with Project MinE or the review.

For the *NEFH* variants identified from the systematic review, 16 out of 59 (27.1%) were found in Project MinE, with 11 of these occurring in Project MinE cases and controls, review and gnomAD (Figure 5c). Examination of case-control frequencies of review-identified variants present in Project MinE (Table 3) suggested that K790del could be protective against ALS (0.139% cases, 0.04% controls; Beta (SE) = -1.03 (0.47), p=0.03). Using the Project MinE cohort as an additional study for meta-analysis of individual variants did not offer any other insights into their contribution to ALS risk (Supplementary Table 6).

**Table 3.**
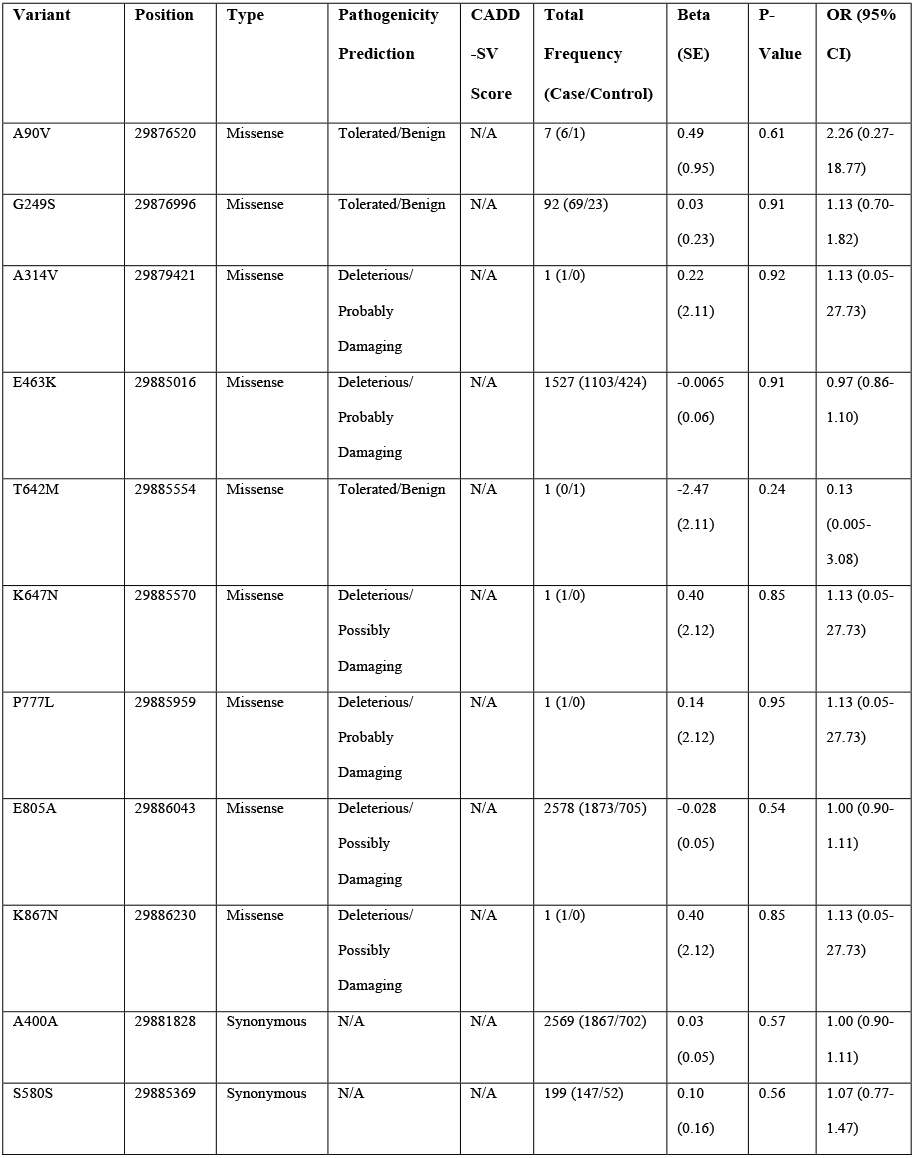
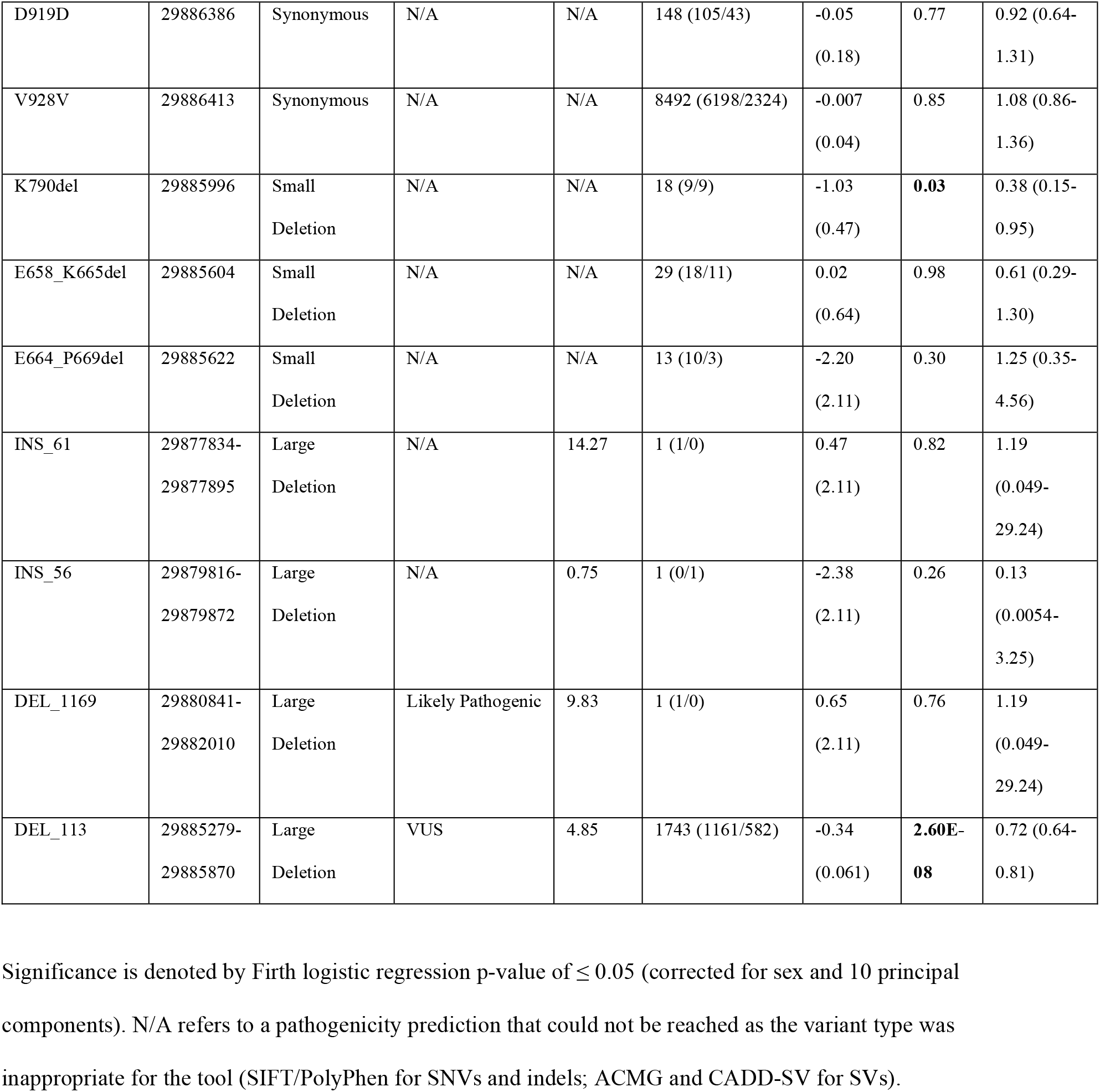
Case-control frequencies of the 16 SNV/indel variants found in both the systematic review and in the Project MinE cohort, and the 4 structural variants identified in a subset of Project MinE.

### Screening of *NEFH* structural variants in the Project MinE cohort

Only 4 structural variants were identified in a subset of the Project MinE cohort (Table 3). All were located in the KSP and KEP segments of the tail domain, with none being likely pathogenic according to the CADD-SV pathogenicity prediction tool (≥ 15). When comparing case-control frequencies to elucidate the association of the structural variants with ALS, the 113bp KEP segment deletion was found to be protective against ALS (17.95% cases vs 23.91% controls; Beta (SE) = -0.34 (0.061), p=2.60E-08).

### Rare variant burden analysis in the Project MinE cohort

All of the SNV/indel variants from Project MinE were subject to burden analysis at two frequency levels (Ultra-Rare (<0.1%) and High-frequency rare (0.1-1%)) to assess the contribution of different variant classes to ALS risk (Supplementary Tables 7 and 8) stratified by domain. We found that ultra-rare pathogenic missense tail variants (PolyPhen and/or SIFT) were associated with an increased risk of developing ALS (OR 1.94, 95% CI 0.86-4.37; Madsen Browning p=0.039), which replicated and confirmed the result of the meta-analysis. Stratifying this by subdomain revealed that the KEP repeat drove this result (OR 5.65, 95% CI 0.75-42.83, Madsen Browning p=0.02), and that other domains mildly increased ALS risk albeit insignificantly (Supplementary Table 7). In line with previous reports, ultra-rare tail domain in-frame deletions had a large impact on ALS risk, but this finding is at the border of the significance testing threshold (OR 3.01, 95% CI 0.69-13.12, Madsen-Browning p=0.052). A similar but significant effect was observed for low-frequency rare in-frame deletions (Supplementary Table 8), with an OR of 1.18 (95% CI 0.67-2.07, SKAT-O p=0.03). Ultra-rare pathogenic missense variants and high-frequency rare in-frame deletions identified and assessed in these burden analyses are detailed in Figure 3 (bottom panel), with their genomic coordinates, variant frequencies and pathogenicity status detailed in Supplementary Table 9.

When assessing the role of moderate impact variants (missense and indel variants, as defined by VEP), both ultra and high-frequency rare tail-domain variants increased ALS risk (Ultra-Rare OR 1.69, 95% CI 1.00-2.87, Madsen-Browning p=0.024; High-Frequency Rare OR 1.13, 95% CI 0.70-1.84, SKAT-O p=0.04), as did moderate impact variants throughout the whole gene (OR 1.47, 95% CI 0.98-2.22, Madsen-Browning p=0.032). Interestingly, ultra-rare intronic, 5’UTR and modifying impact variants (intronic, 5’UTR and 3’UTR combined) also significantly increased ALS risk (Supplementary Table 7).

## DISCUSSION

In this study, we found that missense tail variants in *NEFH* are associated with an increased risk of ALS. The meta-analysis of 3 previous case-control reports which documented rare (MAF<1%) missense tail domain *NEFH* variant frequencies in a total of 1164 ALS patients and 2,177 controls yielded an OR of 4.56 (p<0.0001). This association was replicated, although with a lower OR, when performing an ultra-rare variant burden analysis of pathogenic missense tail variants in the Project MinE dataset (OR 1.94, Madsen-Browning p=0.039). This is likely due to the discrepancy in sample size between the two cohorts, as Project MinE contains more than 5.5 times the number of cases used in the meta-analysis, with smaller sample sizes often reporting a larger effect size (OR) for significant relationships in either direction ^53^ and also the ‘winner’s curse’ effect commonly observed in genetic association discovery studies ^54^.These findings hold high validity as the vast majority of variants in the meta-analysis were deleterious and possibly/probably damaging according to SIFT and PolyPhen pathogenic prediction tools (Supplementary Table 2), which were the same criteria used for missense variants in the burden analysis to be considered pathogenic. Furthermore, removing the tail domain variants from the rare missense variant meta-analysis, which does not take domain-specific effects into account, as well as performing ultra-rare burden analysis of pathogenic missense variants in the whole gene and head and rod domain, nullified the association with ALS, thus proving that missense tail variants were drivers of ALS risk. Additionally, we found that pathogenic ultra-rare missense variants in the KEP repeat were the main drivers of the association with ALS risk in Project MinE (OR 5.65, Madsen Browning p=0.02). However, pathogenic ultra-rare missense variants in the KSP repeat still showed a the same direction of effect in the Project MinE burden analysis.

Unlike its *NEFM* and *NEFL* counterparts, the mechanism by which missense tail variants alter the functionality of the *NEFH* gene has not been fully elucidated ^6^. Despite this, it is plausible to suggest that these mutations, especially those in the KSP repeat segment, could modify the effects of phosphorylation, thereby changing the conformation of the NF-H subunit in such a way that simultaneously increases the propensity of pNF-H aggregate formation in the axon and disrupts energy metabolism and protein transport. Indeed, *NEFH* also has a complex mRNA and protein-linked stoichiometry of which has not been studied here and could provide us with additional insights into the genetic basis of NF-H inclusion formation. For instance, downregulation of two exclusively spinal cord and CSF-expressed miRNAs, miR-92a-3p, miR-9-5p, bind to recognition elements in the 3’UTR of *NEFH* to increase the expression of NEFH mRNA transcripts and NF-H protein levels in people with ALS ^55,56^. Moreover, there is emerging evidence which suggests that microglial-secreted protein factors can influence NEFH transcript expression and contribute to NF-H inclusion pathology, albeit in the absence of *NEFH* mutations ^57^. Therefore, future studies should build on what we have reported here by incorporating genetic evidence of missense tail mutations with proteomic and transcriptomic data to determine if the aberrant stoichiometry of NF-H is due solely to the action of the mutation on phosphorylation sites within the tail or is a product of a larger interaction between miRNA, protein and glial targets.

We also found that high-frequency rare (MAF 0.1-1%) small in-frame deletions in the tail domain confer susceptibility to ALS within Project MinE (OR 1.18, SKAT-O p=0.03), which agrees with previous findings in the literature ^16,17^. However, the literature based meta-analysis did not find a significant association with ALS risk for either rare (MAF<1%, OR 0.90, p=0.85) or ultra-rare (MAF<0.1%, OR 0.94, p=0.93) tail deletions (Table 2). This discrepancy could again be due to the relatively small sample sizes used in the meta-analysis compared to in Project MinE, or that there may be subdomain-specific effects occurring in the tail that the meta-analysis design could not account for. Potentially, deletions in the KSP repeat could be associated with an increased risk for ALS and that perhaps deletions in the KEP segment may dilute this association having a protective effect. This is plausible given that we found a novel protective 113bp deletion in the KEP region (p=2.60E-08), present in 18% of cases and 24% controls. Another possibility is that the rarity of these deletions coupled with the small sample sizes and genetic technologies of studies used in the meta-analysis resulted in decreased power to detect any significant associations or increases in ALS risk. This was the case for several ultra-rare variant categories which were examined using both meta-analysis and variant burden methods, where there were either higher ORs i.e. in-frame deletions, or lower ORs i.e. missense head, missense rod, present in the Project MinE burden analysis. An unexpected finding was that the presence of ultra-rare (MAF<0.1%) intronic rod variants were significantly associated with increased ALS risk (Supplementary Table 7, OR 1.40, Madsen Browning p=0.004), as the rod domain is highly conserved across all of the neurofilament subunit family ^4^, and there has been no evidence to date to support the role of non-coding DNA in neurofilaments to neurodegeneration. Further research on this is needed to establish the role of non-coding neurofilament variants in ALS.

In conclusion, we showed that missense mutations and in-frame deletions in the tail domain of *NEFH* are associated with the increase of ALS risk, using a two-tiered meta-analysis and variant burden approach which leveraged *NEFH* variant information of 11,130 ALS patients and 7,416 controls from both the literature and the largest whole-genome sequencing consortium for ALS, Project MinE. Our results support the inclusion of missense variants and in-frame deletions in the tail of *NEFH* in ALS sequencing panels.

## Data Availability

All data produced in the present study are available upon reasonable request to the authors.

## ACKNOWLEDGEMENTS

The authors are supported by South London and Maudsley NHS Foundation Trust; MND Scotland; Motor Neurone Disease Association; National Institute for Health Research; Darby Rimmer MND Foundation; Spastic Paraplegia Foundation and Rosetrees Trust. H.M is supported by GlaxoSmithKline and the KCL funded centre for Doctoral Training (CDT) in Data-Driven Health. A.A.K is funded by ALS Association Milton Safenowitz Research Fellowship (grant number22-PDF-609.DOI :10.52546/pc.gr.150909.), The Motor Neurone Disease Association (MNDA) Fellowship (Al Khleifat/Oct21/975-799), The Darby Rimmer Foundation, and The NIHR Maudsley Biomedical Research Centre. A.I is funded by the Motor Neurone Disease Association and The NIHR Maudsley Biomedical Research Centre. A.A-C is an NIHR Senior Investigator (NIHR202421) and has received support from an EU Joint Programme - Neurodegenerative Disease Research (JPND) project. The work is supported through the following funding organisations under the aegis of JPND - www.jpnd.eu *(United Kingdom, Medical Research Council* (MR/L501529/1; MR/R024804/1) *and Economic and Social Research Council* (ES/L008238/1)) and through the Motor Neurone Disease Association, My Name’5 Doddie Foundation, and Alan Davidson Foundation. This study represents independent research part funded by the National Institute for Health Research (NIHR) Biomedical Research Centre at South London and Maudsley NHS Foundation Trust and King’s College London. The authors acknowledge use of the research computing facility at King’s College London, Rosalind (https://rosalind.kcl.ac.uk). The views expressed are those of the author(s) and not necessarily those of the NHS, the NIHR or the Department of Health and Social Care. Project MinE Belgium was supported by a grant from IWT (n° 140935), the ALS Liga België, the National Lottery of Belgium and the KU Leuven Opening the Future Fund. PVD holds a senior clinical investigatorship of FWO-Vlaanderen (G077121N) and is supported by the E. von Behring Chair for Neuromuscular and Neurodegenerative Disorders, the ALS Liga België and the KU Leuven funds “Een Hart voor ALS”, “Laeversfonds voor ALS Onderzoek” and the “Valéry Perrier Race against ALS Fund”. Several authors of this publication are member of the European Reference Network for Rare Neuromuscular Diseases (ERN-NMD)

## AUTHOR CONTRIBUTIONS

H.M, A.A-C and A.I contributed to conception and design of the study; H.M, T.P.S, A.A.K and A.I contributed to the acquisition and analysis of data; all authors contributed to drafting the text or preparing the figures. Details about the contributing members of the Project MinE ALS Sequencing Consortium and their affiliated institutions are available in Supplementary Table 10.

